# Early dynamics of transmission and control of COVID-19: a mathematical modelling study

**DOI:** 10.1101/2020.01.31.20019901

**Authors:** Adam J Kucharski, Timothy W Russell, Charlie Diamond, Yang Liu, CMMID nCoV working group, John Edmunds, Sebastian Funk, Rosalind M Eggo

## Abstract

**Background:** An outbreak of the novel coronavirus SARS-CoV-2 has led to 46,997 confirmed cases as of 13^th^ February 2020. Understanding the early transmission dynamics of the infection and evaluating the effectiveness of control measures is crucial for assessing the potential for sustained transmission to occur in new areas.

**Methods:** We combined a stochastic transmission model with data on cases of novel coronavirus disease (COVID-19) in Wuhan and international cases that originated in Wuhan to estimate how transmission had varied over time during January and February 2020. Based on these estimates, we then calculated the probability that newly introduced cases might generate outbreaks in other areas.

**Findings:** We estimated that the median daily reproduction number, *R*_*t*_, declined from 2.35 (95% CI: 1.15-4.77) one week before travel restrictions were introduced on 23^rd^ January to 1.05 (95% CI: 0.413-2.39) one week after. Based on our estimates of *R*_*t*_,we calculated that in locations with similar transmission potential as Wuhan in early January, once there are at least four independently introduced cases, there is a more than 50% chance the infection will establish within that population.

**Interpretation:** Our results show that COVID-19 transmission likely declined in Wuhan during late January 2020, coinciding with the introduction of control measures. As more cases arrive in international locations with similar transmission potential to Wuhan pre-control, it is likely many chains of transmission will fail to establish initially, but may still cause new outbreaks eventually.

**Funding:** Wellcome Trust (206250/Z/17/Z, 210758/Z/18/Z), HDR UK (MR/S003975/1), Gates Foundation (INV-003174), NIHR (16/137/109)

## Introduction

As of 13^th^ February 2020, an outbreak of COVID-19 has resulted in 46,997 confirmed cases (1). The outbreak was first identified in Wuhan, China, in December 2019, with the majority of early cases being reported in the city. The majority of internationally exported cases reported to date have a travel history to Wuhan (2). In the early stages of a new infectious disease outbreak, it is crucial to understand the transmission dynamics of the infection. In particular, estimation of changes in transmission over time can provide insights into the current epidemiological situation (3) and identify whether outbreak control measures are having a measurable effect (4,5). Such analysis can inform predictions about potential future growth (6), help estimate risk to other countries (7), and guide the design of alternative interventions (8).

There are several challenges to such analysis, however, particularly in real-time. There can be a delay to symptom appearance resulting from the incubation period and delay to confirmation of cases resulting from detection and testing capacity (9). Modelling approaches can account for such delays and uncertainty, by explicitly incorporating delays resulting from the natural history of infection and reporting processes (10). In addition, individual data sources may be biased, incomplete, or only capture certain aspects of the outbreak dynamics. Evidence synthesis approaches, which fit to multiple data sources rather than a single dataset (or data point) can enable more robust estimation of the underlying dynamics of transmission from noisy data (11,12). Combining a mathematical model of SARS-CoV-2 transmission with four datasets from within and outside Wuhan, we estimated how transmission in Wuhan varied between December and February 2020. We then used these estimates to assess the potential for sustained human-to-human transmission to occur in locations outside Wuhan if cases are introduced.

## Research in Context

### Evidence before this stud

We searched PubMed, BioRxiv and MedRxiv for articles published up to 10^th^ February 2020 using the keywords “2019-nCoV”, “novel coronavirus”, “COVID-19”, “SARS-CoV-2” AND “reproduction number”, “R0”, “transmission”. We found several estimates of the basic reproduction number, R_0_, including average exponential growth rate estimates based on inferred or observed cases at a recent time point (13,14) and early growth of the outbreak in China (15,16). However, we identified no estimates of how R_0_ had changed in Wuhan since control measures were introduced in late January, or estimates that jointly fitted to data within Wuhan with international exported cases and evacuation flights.

### Added value of this study

Our study combines available evidence from multiple data sources, reducing the dependency of our estimates on a single time point or dataset. We estimate how transmission has varied over time, identify a decline in the reproduction number in late January to near 1, coinciding with the introduction of large scale control measures, and show the potential implications of estimated transmission for outbreak risk new locations.

### Implications of all the available evidence

COVID-19 is currently exhibiting sustained transmission in China. This creates a substantial risk of outbreaks in other countries, although if SARS-CoV-2 has MERS-CoV or SARS-CoV-like variability in transmission at the individual-level, multiple introductions may be required before an outbreak takes hold.

## Methods

To estimate the early dynamics of transmission in Wuhan, we fitted a stochastic transmission dynamic model (17) to multiple publicly available datasets on cases in Wuhan and internationally exported cases from Wuhan. The four datasets we fitted to were: daily number of new internationally exported cases (or lack thereof), by date of onset, as of 26th January 2020; daily number of new cases in Wuhan with no market exposure, by date of onset, between 1st December 2019 and 1st January 2020; daily number of new cases in China, by date of onset, between 29th December 2019 and 23^rd^ January 2020; proportion of infected passengers on evacuation flights between 29^th^ January and 4^th^ February 2020. We used an additional two datasets for comparison with model outputs: daily number of new exported cases from Wuhan (or lack thereof) in countries with high connectivity to Wuhan (i.e. top 20 most at risk), by date of confirmation, as of 10th February 2020; data on new confirmed cases reported in Wuhan between 16^th^ January and 11^th^ February 2020 (full details in the Appendix).

In the model, individuals were divided into four infection classes (Figure 1): susceptible, exposed (but not yet infectious), infectious, and removed (i.e. isolated, recovered or otherwise no longer infectious). The model accounted for delays in symptom onset and reporting, as well as uncertainty in case observation (see Appendix for full model details). The incubation period was assumed to be Erlang distributed with mean 5.2 days (16) and delay from onset-to-isolation Erlang distributed with mean 2.9 days (2,15). The delay from onset-to-reporting was assumed to be exponentially distributed with mean 6.1 days (2). Once exposed to infection, a proportion of individuals travelled internationally and we assumed that the probability of cases being exported from Wuhan to a specific other country depended on the number of cases in Wuhan, the number of outbound travellers (assumed to be 3300 per day before travel restrictions were introduced on 23rd January 2020 and zero after), the relative connectivity of different countries (18), and the relative probability of reporting a case outside Wuhan, to account for differences in clinical case definition, detection and reporting within Wuhan and internationally. We considered the 20 countries outside China most at risk of exported cases in the analysis.

**Figure 1:**
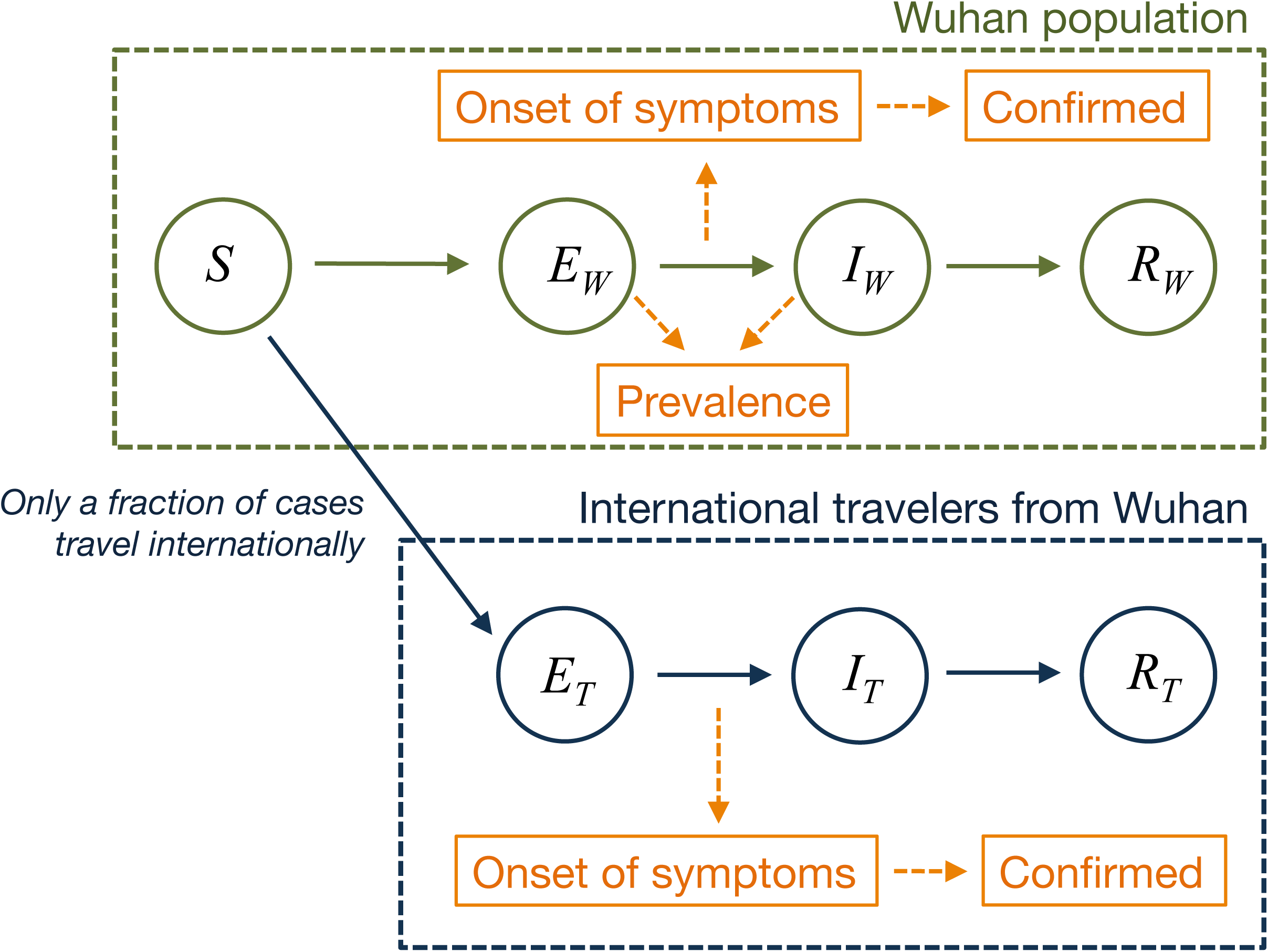
Model structure. The population is divided into four classes: susceptible, exposed (and not yet symptomatic), infectious (and symptomatic), removed (i.e. isolated, recovered, or otherwise non-infectious). A fraction of exposed individuals subsequently travel and are eventually detected in their destination country.

Transmission was modelled as a geometric random walk process, and we used sequential Monte Carlo to infer the transmission rate over time, as well as the resulting number of cases and the time-varying basic reproduction number, *R*_*t*_, defined here as the average number of secondary cases generated by a typical infectious individual on each day in a full susceptible population. The model had three unknown parameters, which we estimated: magnitude of temporal variability in transmission, proportion of cases that would eventually be detectable, and relative probability of reporting a confirmed case within Wuhan compared to an internationally exported case originated in Wuhan. We assumed the outbreak started with a single infectious case on 22^nd^ November 2019 and the entire population was initially susceptible. Once we had estimated *R*_*t*_, we used a branching process with a negative binomial offspring distribution to calculate the probability an introduced case would cause a large outbreak. We also conducted sensitivity analysis on assumptions about the initial number of cases, connectivity between countries and proportion of cases that were infectious before showing symptoms. More details of methodology, sensitivity analysis, data and code availability are provided in the Appendix.

## Results

We estimated that the daily reproduction number, *R*_*t*_, varied during January 2020, with median values ranging from 1.6–2.6 between 1st January 2020 and the introduction of travel restrictions on 23rd January (Figure 2A). We estimated a decline in R_t_ in late January, from 2.35 (95% CI: 1.15-4.77) on 16^th^ of January, one week before the restrictions, to 1.05 (95% CI: 0.413-2.39) on 31^st^ January.

**Figure 2:**
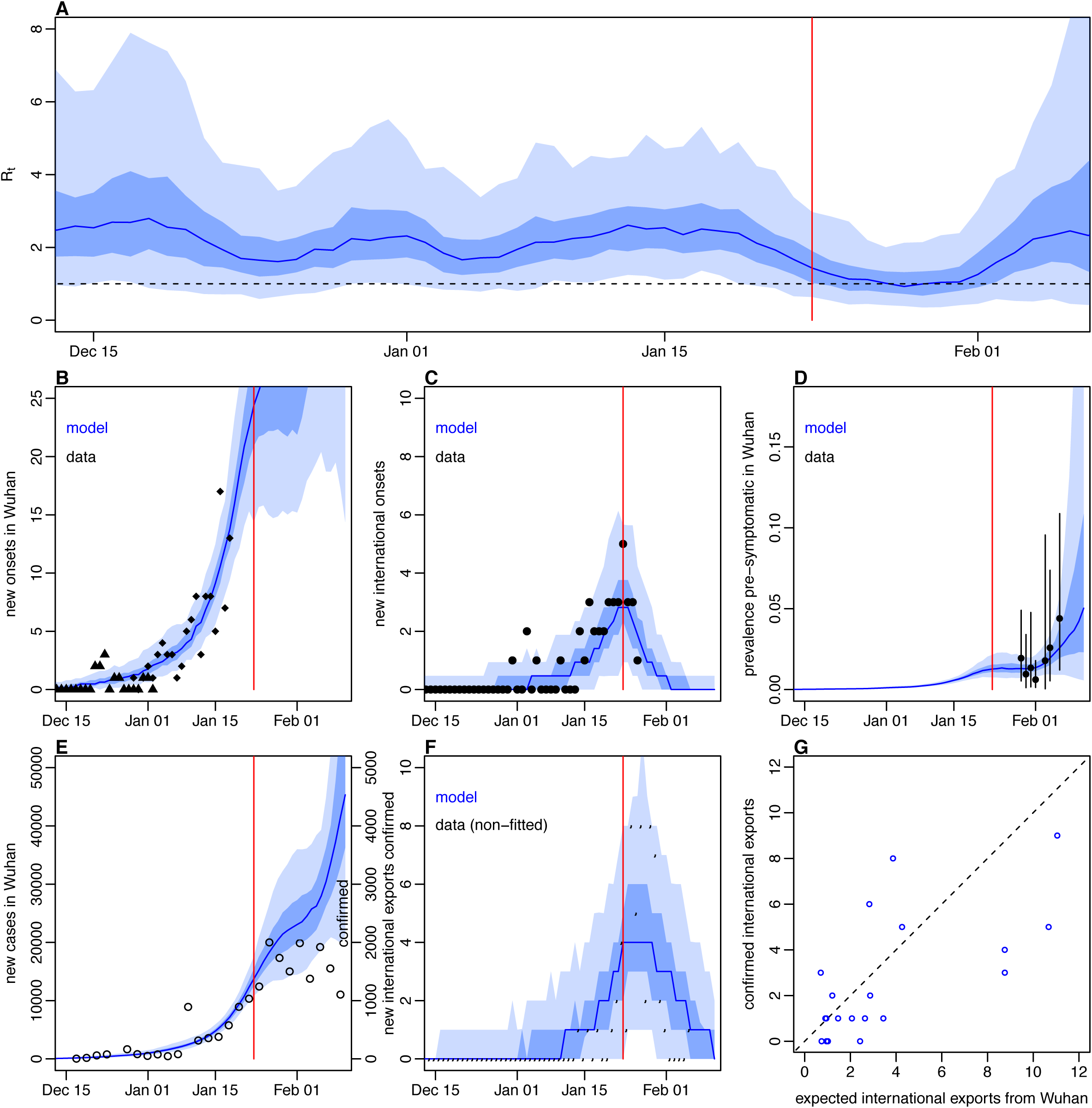
Dynamics of transmission in Wuhan, fitted up to 13th February 2020. Red line marks travel restrictions starting on 23^rd^ January 2020. A) Estimated daily reproduction number (R_t_) over time. B) Onset dates of confirmed cases in Wuhan (triangles) and China (diamonds). Blue lines and shaded regions: median, 50% and 95% credible intervals of model estimate. C) Reported cases by date of onset (black) and estimated internationally exported cases from Wuhan by date of onset (blue line). D) Estimated prevalnece of infections that do not have detectable symptoms (blue line), and proportion of passengers on evacuation flights that tested positive for SARS-CoV-2 (black points, with 95% binomial CIs shown). E) New confirmed cases by date in Wuhan (circles, right hand axis) and estimated new symptomatic cases (blue line, left hand axis). F) International exportation events by date of confirmation of case, and expected number of exports in the fitted model. G) Estimated number of internationally exported from Wuhan confirmed up to 10^th^ Feburary 2020 and observed number in 20 countries with highest connectivity to China. In all panels, datasets that were fitted to shown as solid points; non-fitted data shown as circles.

The model reproduced the observed temporal trend of cases within Wuhan and cases exported internationally, capturing all of the dynamics reflected by these different data streams (Figure 2B–D). Our results suggested there were around tenfold more symptomatic cases in Wuhan in late January than were reported as confirmed cases (Figure 2E), but the model not predict the slowdown in cases that was observed in early February. The model could also reproduce the pattern of confirmed exported cases from Wuhan, which was not explicitly used in the model fitting (Figure 2F). We found that confirmed and estimated exported cases among the twenty countries most connected to China were generally in good correspondence, with the USA and Australia as notable outliers, having had more confirmed cases reported with a travel history to Wuhan than would be expected in the model (Figure 2G). We estimated that 100% (95% CI: 51–100%) of cases would be eventually had detectable symptoms, implying that most infections that were exported internationally from Wuhan in late January were eventually detected.

To examine the potential for new outbreaks to establish in locations outside of Wuhan, we used our estimates of the reproduction number to simulate new outbreaks with potential individual-level variation in transmission (i.e. ‘superspreading events’) (13,19,20). Such variation increases the fragility of transmission chains, making it less likely that an outbreak will take off following a single introduction; if transmission is more homogeneous, with all infectious individuals generating a similar number of secondary cases, it is more likely than an outbreak will establish (19). Based on the median reproduction number estimated during January before travel restrictions were introduced, we estimated that a single introduction of 2019-nCoV with SARS-like or MERS-like individual-level variation in transmission would have a 20–28% probability of causing a large outbreak (Figure 3A). Assuming SARS-like variation and Wuhan-like transmission, we estimated that once four or more infections have been introduced into a new location, there is an over 50% chance that an outbreak will occur (Figure 3B).

**Figure 3:**
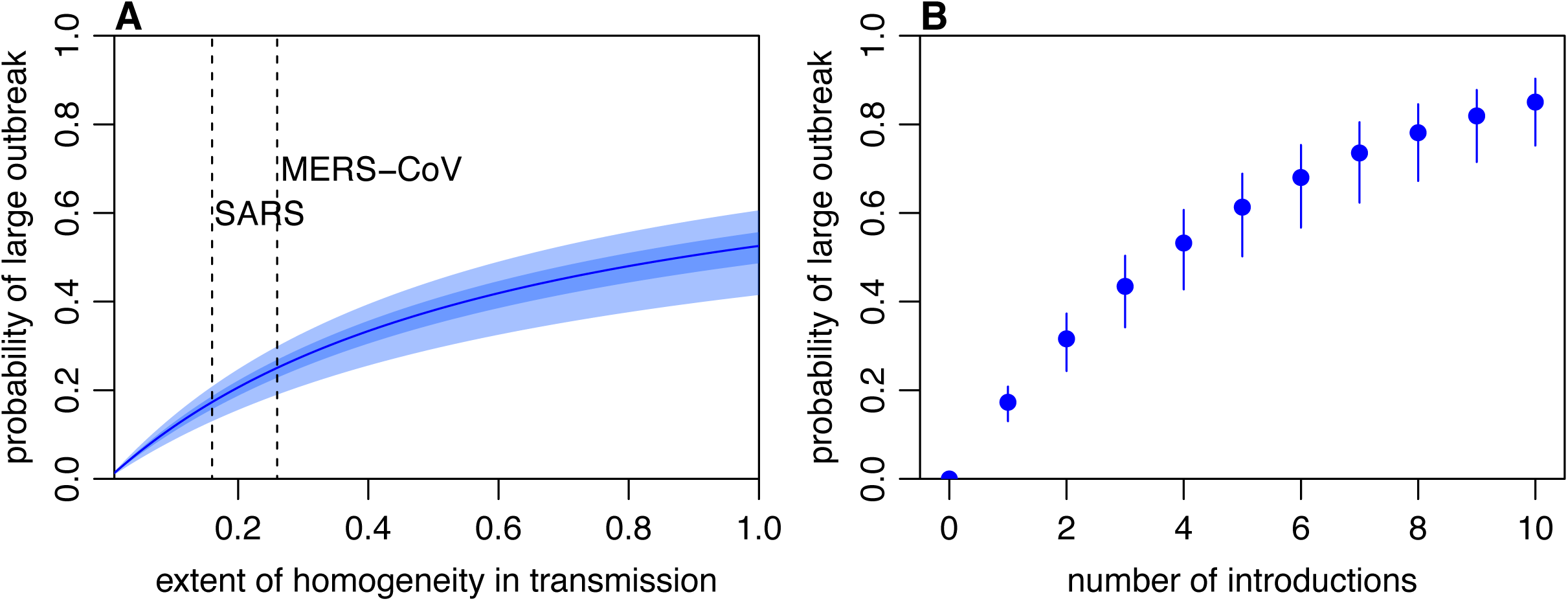
Risk that introduced infections will establish in a new population. A) Probability a single case will lead to a large outbreak for different assumptions about the extent of homogeneity in individual-level transmission (i.e. the dispersion parameter k in a negative binomial offspring process). Results are shown for the median reproduction number estimated for nCoV-2019 in Wuhan between 1st January and 23rd January 2020 and. B) Probability a given number of introductions will result in a large outbreak, assuming SARS-like superspreading events can occur.

## Discussion

Combining a mathematical model with multiple datasets, we found that the median daily reproduction number, *R*_*t*_, of SARS-CoV-2 in Wuhan likely varied between 1.6– 2.6 in January 2020 prior to travel restrictions being introduced. We also estimated that transmission declined by around half in the two weeks spanning the introduction of restrictions.

The estimated fluctuations in *R*_*t*_ were driven by the rise and fall in number of cases both in Wuhan and internationally, as well as prevalence on evacuation flights (Figures 2B–D). Such fluctuations could be the result of changes in behaviour in the population at risk, or specific superspreading events that inflated the average estimate of transmission (13,19,20). We found some evidence of reduction in *R*_*t*_ in the days prior to the introduction of travel restrictions in Wuhan, which may have been reflected outbreak control efforts or growing awareness of SARS-CoV-2 during this period. The uncertainty in our estimates for R_t_ following the decline in early February (Figure 2A) results from limited data sources to inform changes in transmission during this period.

Comparing model predictions to observed confirmed cases reported in Wuhan during late January and early February, we found that the model predicts ten-fold higher cases than have been reported; the model also does not predict the recent slowdown in cases, suggesting a potential change in reporting rather than a genuine slowdown in transmission in early February. Our estimates for international cases in specific countries were broadly consistent with the number of subsequently confirmed exported cases outside of Wuhan. However, there were notably more cases exported to France, US, and Australia compared to what our model predicted. This may be the result of increased surveillance and detected as awareness of SARS-CoV-2 grew in late January, which would suggest earlier exported cases may have missed; it may also be the result of increased travel out of Wuhan immediately prior to travel restrictions being introduced on 23rd January.

Based our on estimated reproduction number, and published estimates of individual-level variation in transmission for SARS and MERS-CoV, we found that a single case introduced to a new location would not necessarily lead to an outbreak. Even if the reproduction number is as high as it has been in Wuhan in early January, it may take several introductions for an outbreak to establish; this is because high individual-level variation in transmission makes new chains of transmission more fragile, and hence less likely that a single infection will generate out outbreak. This highlights the importance of rapid case identification, and subsequent isolation and other control measures to reduce the chance of onward chains of transmission (21).

Our analysis highlights the value of combining multiple data sources in analysis of COVID-19. For example, the rapid growth of confirmed cases globally during late January 2020, with case totals in some instances apparently doubling every day or so, would have had the effect of inflating *R*_*t*_ estimates to implausibly large values if only these recent data points were used in analysis. Our results also have implications for the estimation of transmission dynamics using the number of exported cases from a specific area (22). Once extensive restrictions are introduced, as they were in Wuhan, the signal from such data gets substantially weaker. If restrictions and subsequent delays in detection of cases is not accounted for, it could lead to artificially low estimates of *R*_*t*_ or inferred case totals from the apparently declining numbers of exported cases. Our model estimates benefitted from the availability of testing data from evacuation flights, which allowed us to estimate current prevalence. Having such information for other settings, either through widespread testing or serological surveillance, will be valuable to reduce reliance on case reports alone.

There are several other limitations to our analysis. We used plausible biological parameters for SARS-CoV-2 based on current evidence, but these values may be refined as more comprehensive data become available. However, by fitting to multiple datasets to infer model parameters, and performing sensitivity analyses on key areas of uncertainty, we have attempted to make the best possible use of the available evidence about SARS-CoV-2 transmission dynamics. Further, we used publicly available connectivity and risk estimates based on international travel data to predict the number of exported cases into each country. These estimates have shown good correspondence with the distribution of exported cases to date (23), and are similar to another risk assessment for COVID-19 with different data (24). We also assumed that the latent period is equal to the incubation period (i.e. individuals become infectious and symptomatic at the same time) and all infected individuals will eventually become symptomatic. However, there is evidence that transmission of SARS-CoV-2 can occur with limited reported symptoms (25). We therefore conducted a sensitivity analysis in which transmission could occur in the second half of the incubation period, but this did not change our overall conclusions (Appendix, page 7). We also explored having a larger initial spillover event and also using different sources for flight connectivity data, neither of which changed the conclusions of the analysis. In our analysis of new outbreaks, we also used estimates of individual-level variation in transmission for SARS and MERS-CoV to illustrate potential dynamics. However, it remains unclear what the precise extent of such variation is for SARS-CoV-2 (13); if transmission were more homogenous than SARS of MERS-CoV, it would increase the risk of outbreaks following introduced cases. As more data becomes available, it will be possible to refine these estimates, and therefore we made an online tool so users can explore these risk estimates if new data become available (Appendix, page 4)

Our results demonstrate that there was likely substantial variation in SARS-CoV-2 transmission over time, and suggest a decline in transmission in Wuhan in late January around the time that control measures were introduced. If COVID-19 transmission establishes outside of Wuhan, understanding the effectiveness of control measures in different settings will be crucial for understanding the likely dynamics of the outbreak, and the likelihood that transmission can eventually be contained.

## Data Availability

All data and code are available at: https://github.com/adamkucharski/2020-ncov/stoch_model_V2_paper

## Author Contributions

Data analysis was led by AJK, who programmed the model with help from TWR. AJK, SF, and RME planned the inference framework. CD provided the data from online sources. The following authors were part of the CMMID 2019-nCoV working group: Fiona Sun, Mark Jit, James D Munday, Nicholas Davies, Amy Gimma, Kevin van Zandvoort, Hamish Gibbs, Joel Hellewell, Christopher I Jarvis, Sam Clifford, Billy J Quilty, Nikos I Bosse, Sam Abbott, Petra Klepac and Stefan Flasche. Each contributed in processing, cleaning and interpretation of data, interpreted findings, contributed to the manuscript, and approved the work for publication. All authors interpreted the findings, contributed to writing the manuscript and approved the final version for submission.

## Acknowledgements

We would like to thank Motoi Suzuki for his help in identifying the flight evacuation data sources.

